# Actionability of Synthetic Data in a Heterogeneous and Rare Healthcare Demographic; Adolescents and Young Adults (AYAs) with Cancer

**DOI:** 10.1101/2024.03.04.24303526

**Authors:** J. Joshi Hogenboom, A. Aiara Lobo Gomes, A.L.A.J. Andre Dekker, W.T.A. Winette Van Der Graaf, O. Olga Husson, L.Y.L. Leonard Wee

## Abstract

**Purpose:** Research on rare diseases and atypical healthcare demographics is often slowed by high inter-subject heterogeneity and overall scarcity of data. Synthetic data (SD) has been proposed as means for data sharing, enlargement, and diversification, by artificially generating ‘real’ phenomena while obscuring the ‘real’ subject data. The utility of SD is actively scrutinised in healthcare research, but the role of sample size for actionability of SD is insufficiently explored. We aim to understand the interplay of actionability and sample size by generating SD sets of varying sizes from gradually diminishing amounts of real subjects’ data. We evaluate the actionability of SD in a highly heterogeneous and rare demographic: adolescents and young adults (AYAs) with cancer.

**Methodology:** A population-based cross-sectional cohort study of 3735 AYAs was sub-sampled at random to produce 13 training datasets of varying sample sizes. We studied four distinct generator architectures built on the open-source Synthetic Data Vault library. Each architecture was used to generate SD of varying sizes based on each aforementioned training subsets. SD actionability was assessed by comparing the resulting SD to its respective ‘real’ data against three metrics – veracity, utility, and privacy concealment.

**Results:** All examined generator architectures yielded actionable data when generating SD with sizes similar to the ‘real’ data. Large SD sample size increased veracity but generally increased privacy risks. Using fewer training subjects led to faster convergence in veracity, but partially exacerbated privacy concealment issues.

**Conclusion:** SD is a potentially promising option for data sharing and data augmentation, yet sample size plays a significant role in its actionability. SD generation should go hand-in-hand with consistent scrutiny and sample size should be carefully considered in this process.

## Introduction

Healthcare data for rare diseases in heterogeneous populations – such as adolescents and young adults (AYAs) with cancer – are generally difficult to acquire, and sizable high-quality datasets on these subjects are not widely available. Research on this important group has thus been hampered by data availability, resulting in lack of standards for age-specific cancer care^1^. Sharing of datasets is regularly constrained by administrative burden and privacy-related challenges. However, it is exactly the research into such diseases that would benefit most from greater access to larger volumes of high-quality data.

Synthetic data (SD) has been suggested as a potential surrogate that addresses data availability and data acquisition^2–7^, since it has means to augment both sample size and data diversity. Here, SD is defined as artificially generated person-level health data that resembles – to some degree of authenticity – ‘real’ subject health data, whilst implying the absence of identifiable information of any actual persons. In this case, the goal of SD is to catalyse knowledge generation in healthcare, but without exposing real human subjects’ data. While use of SD within healthcare is still in development, ethicists, policy makers, and researchers are trying to establish how SD can be used appropriately^5,8^. Meanwhile, various techniques^9–12^ are now openly available which researchers can use to train generative models and thus create SD for various purposes^13–22^.

Constructing a SD generator always needs some initial form of real data for its training. A trained generative network (i.e., generator) is then used to produce any arbitrary size of SD. The sample size of training data is therefore an important question that also depends on the intended use of SD. Likewise, it is not universally known – for a given generator – if there are upper and lower sample size limits of SD that ought to be produced. The interplay of actionability and sample size (both for training and generation) therefore warrants careful examination.

What designates SD as “actionable” remains a disputed topic^2,23,24^; in this work, we propose three evaluation domains that may be considered as minimally necessary for actionable SD. (1) Veracity, meaning that the generated SD is statistical similar to the training data in terms of distribution of values. (2) Utility, meaning that SD replicates the inter-variable associations within the training data, for example, the coefficients in a logistic regression model. (3) Privacy concealment, in terms of obscuring individual identity and obscuring their observable attributes.

To elucidate the dependence of veracity, utility, and privacy concealment on sample size, we ran training and SD-generating experiments using real AYA cancer data (persons aged 18 to 39 years at first cancer diagnosis). Cancer incidence is low in this demographic (only 1 million new cases annually worldwide), making this group difficult to study by conventional methods^1^. Though this group experiences the burden of cancer in a starkly different way compared to paediatric and elderly patients^25,26^, AYAs are predominantly treated in either paediatric or adult care institutes. Evidence to inform personalised age-specific patterns of cancer care is very limited for AYAs, hence our interest in understanding how SD might potentially be used to address AYA health-related hypotheses.

## Methods

Data from the SURVAYA study^25,26^ (clinicaltrials.gov NCT05379387) was re-used with permission from the sponsor. SURVAYA was a population-based cross-sectional cohort study amongst AYA cancer patients registered in the Netherlands Cancer Registry (NCR). SURVAYA included AYAs that had been treated either in a Dutch academic medical centre or in the Netherlands Cancer Institute, and were registered in the NCR between 1999 to 2015; Amsterdam AMC, Netherlands Cancer Institute, and Utrecht UMC supplied treatment-related data up to 2014. The principal instruments in SURVAYA were validated health-related Quality of Life (QoL) questionnaires^27–30^, which were linked to clinical attributes from the NCR. Details are provided in the original study publications^25,26^. In total, there were 950 variables, some of which contained unstructured text.

We limited ourselves only to variables selected by Saris et al. (2022) for logistic regression models of negative body image among AYAs^25^. As per the original research, participants that had filled in less than half of the body image-related questionnaire items were excluded. We imputed missing values using multiple imputation by chained equations. Age at diagnosis was left in as a privacy concealment challenge for attribute inference, since certain cancer diagnoses are widely known to be exceptionally rare among younger AYAs. The pre-processed training data, hereafter referred to as the original dataset (OD), consisted of 3735 AYAs with 21 variables (37 after a one-hot encoding of categories) – see Supplementary material S1.

Training and generating experiments are illustrated in Figure 1. The OD was sampled without replacement to produce smaller training datasets of sizes 3600, 3350, …, down to 600 subjects, resulting in a total of 14 distinct training sets.

**Figure 1:**
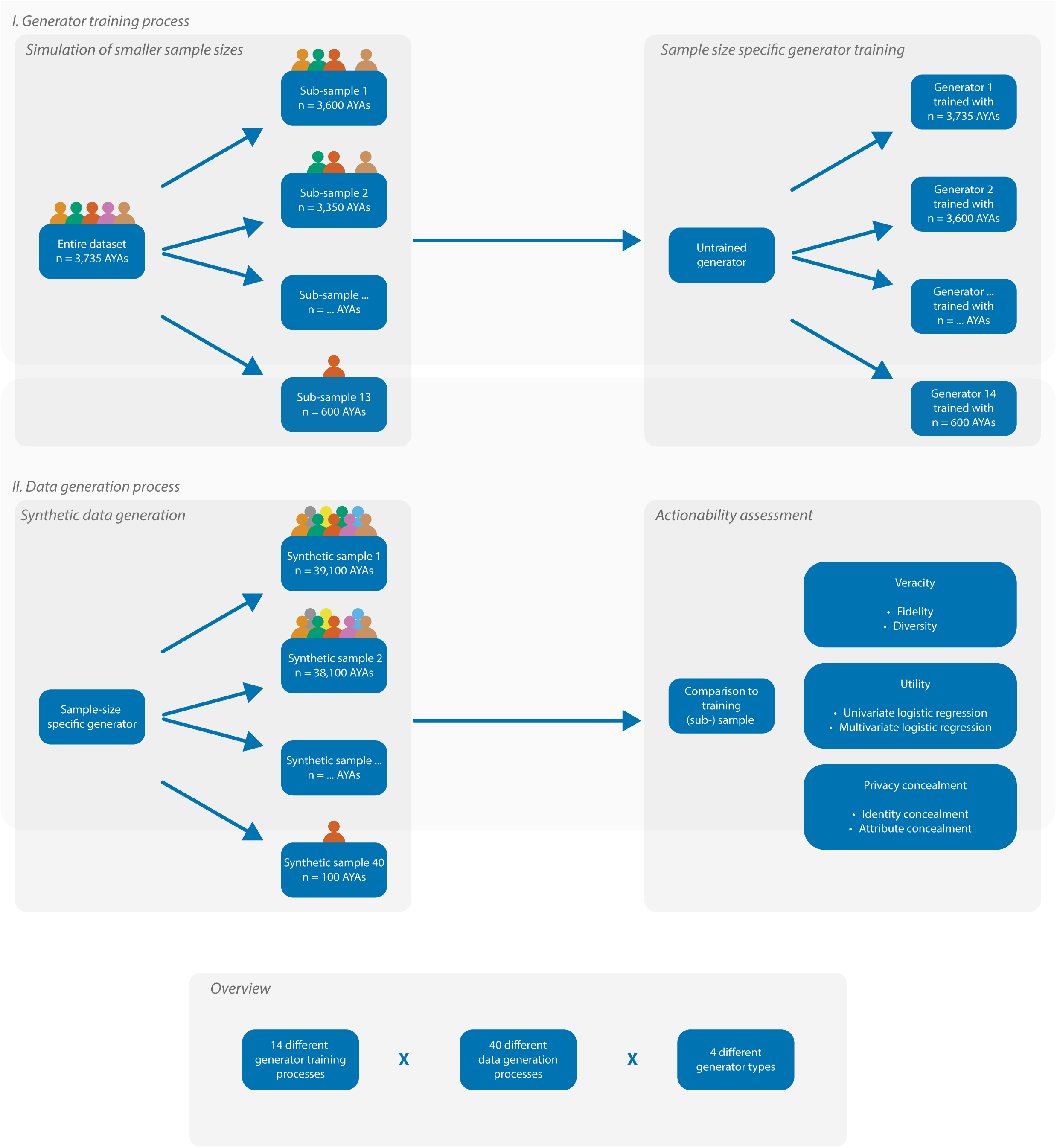
Overview that describes the experimental setup to evaluate the role of OD and SD sample size on SD actionability.

We tested four SD generator architectures from the Synthetic Data Vault (SDV)^9^ Python library. Two of these were machine learning (ML)-based architectures that used strictly parametric statistical models. The first was a speed-optimised Gaussian copula generator (hereafter “Fast ML”) and the other was a classical Gaussian copula generator (hereafter “Gaussian Copula”). Two non-parametric architectures were based on deep-learning (DL) neural networks; a Conditional Tabular Generative Adversarial Network (CTGAN)^12^ and a Conditional Generative Adversarial Network incorporating Differential Privacy (DP-CGAN)^10^. The DP-CGAN was not part of SDV’s current library, but was built on top of existing SDV-library modules.

In the Fast ML architecture, each of the aforementioned subsets of the OD were used for training, thus resulting in 14 separately trained Fast ML generators. This process was then repeated entirely for the Gaussian Copula, CTGAN, and DP-CGAN architectures, thus always resulting in 14 separately trained generators within each of the four selected architectures.

Each architecture used default (hyper-)parameters for training specified by its developers. We limited our experiments only to the default hyper-parameters, without fine-tuning and no grid search for these experiments. All architectures were supplied with the metadata of the OD as required in the documentation^10,31^. Hyper-parameters per architecture and variable metadata are provided in supplementary material S2 and S3, respectively.

From each of the separately trained generators (thus 4 architectures x 14 training OD subsets per architecture = 56 unique generators), we generated SD of varying samples sizes ranging from 100 up to 39100 artificial subjects, in increments of 1000. Thus, for each uniquely trained generator, we derived 40 distinct sets of SD (56 generators x 40 sets of SD per generator = 2240 SD sets in total).

Actionability was assessed by comparing a selected SD set, from a given generator, to the corresponding OD training set used to train that generator. For instance, a SD set of 5100 samples generated with a Fast ML generator that was trained using a set of 3735 real subjects was compared to those specific 3735 real subjects.

Veracity of SD was quantified in terms of precision, density, recall, and coverage^32–34^. The precision metric quantifies the proportion of individual subjects in the SD that fall within a minimum number of neighbouring subjects (k) found in the OD. The density corrects for outliers in precision by scoring the number of samples inside the densely overlapping regions. Recall describes the proportion of OD samples that lay close to the SD’s samples. Finally, coverage corrects for outliers in recall by building the radii with the OD’s samples, rather than the SD’s samples. A schematic overview of the veracity metrics’ mechanism is provided in supplementary material S4. The metrics were computed using the *prdc* Python library with a constant k = 5.

Utility was examined by checking if a given SD reproduced the same logistic regression coefficients as the original paper by Saris et al.^25^, for generative architectures that had been trained using all of 3735 real subjects. For each generator, we selected an SD output size of 3100 for the comparison of odds ratios (ORs) with Saris et al. The regression analyses were conducted using Python libraries; *Statsmodels* for regression analysis and *scikit-learn* for an optimism-adjusted (through random under-sampling) concordance-statistic as area-under receiver-operator curve score.

Privacy concealment was assessed with two metrics. First, a measure of the minimum Hamming distance^10^ between any ‘synthetic’ subject in a given SD to any ‘real’ subject in the OD used to train the corresponding generator. Second, we estimated an attribute inference probability using seemingly innocent information that might be obtained by through public channels. We limited the test variables to age in nearest whole year, type of cancer and romantic partnership status, which we assumed would be easily gleaned from social media. We then assumed that an attacker would infer a sensitive and not easily known attribute, i.e., an AYA’s self-perception of sexual attractiveness (a primary outcome in SURVAYA). The attribute inference attack was simulated using a Correct Attribution Probability algorithm (CAP)^35^.

## Results

### Veracity

Figure 2 shows a sensitivity of coverage towards training and SD sample size. For the generator architectures we tested, reasonable coverage (> 0.75) was obtained for a wide range of training sample sizes, as long as more than 3000 synthetic subjects were generated. Among the four architectures, CTGAN overall yielded lower coverage for combinations of training and SD sample sizes. When the original data was relatively small in size and lacking in diversity, the chance of finding synthetic subjects within the radii of 5 closest real subjects was relatively high, hence the coverage seems counterintuitively better for smaller training set size.

**Figure 2:**
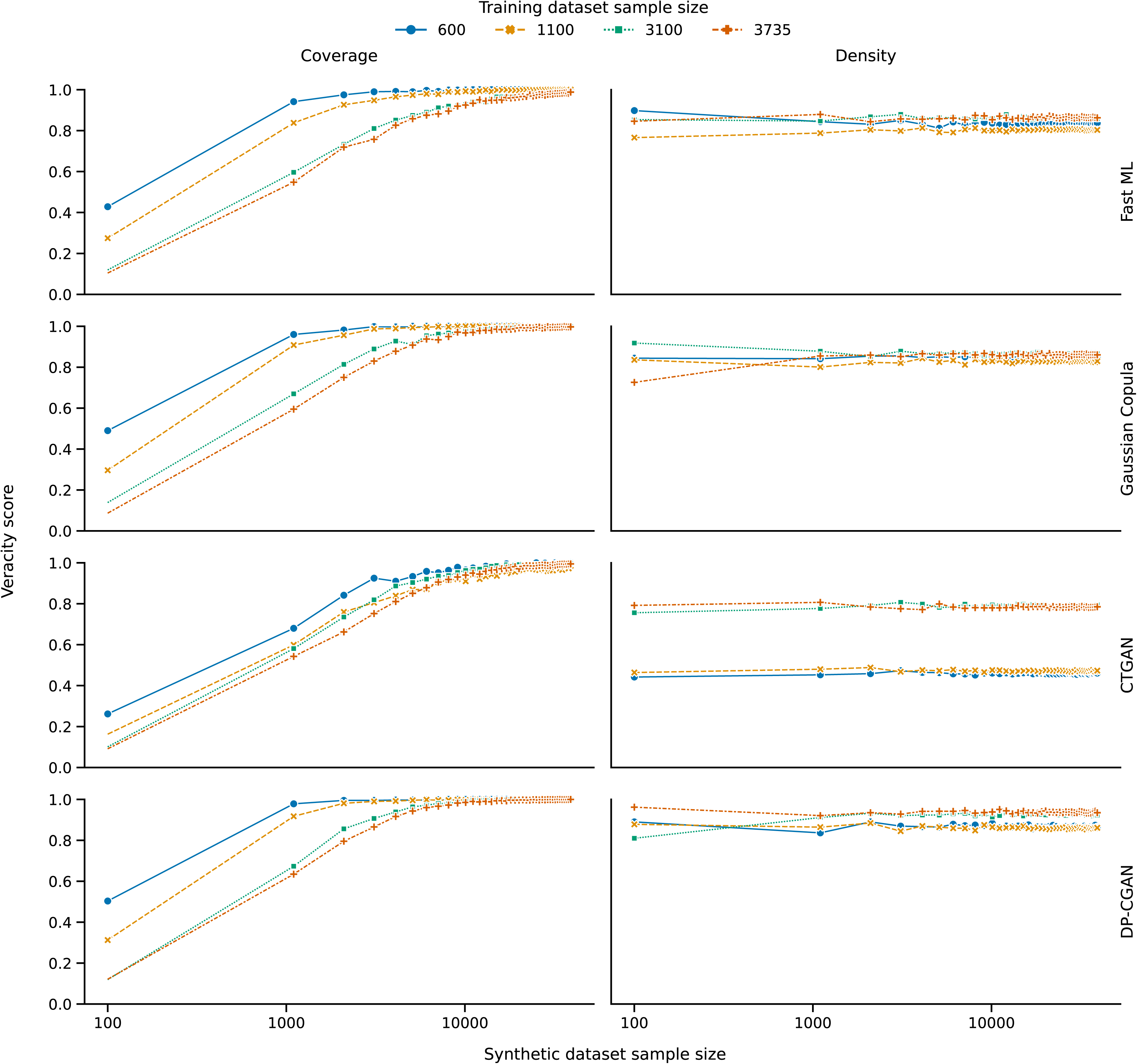
Veracity in terms of coverage (on the left side of the figure) and density (on the right side of the figure), per colour-coded training data sample size. The veracity score relates to the prdc’s outcome, where a value of 1 is best, and 0 is worst. Each horizontal row of figures depict the respective scores of the included generator architectures.

Density was for the most part independent of training and SD sample size, as shown in Figure 2. However, CTGAN shows a stepwise improvement in the density metric from 1100 to 3100 training sample size. Similarly, the precision and recall scores were acceptably high and were independent of either training or SD sample size; the precision and recall curves are provided in supplementary material S5 and minimum scores were 0.86, 0.90, 0.75, and 0.93 for Fast ML, Gaussian Copula, CTGAN, and DP-CGAN respectively for both precision and recall.

### Utility

The overall picture for the ORs in univariable and multivariable logistic regressions against negative body image was quite mixed. There was a moderate degree of overlapping associations among the variables, where the confidence interval of the OR estimated from the SD included the same OR estimated in the OD. However, the number of overlapping associations between the SD and OD varied between different generator architectures.

Further, ORs that were not statistically significant in the OD (p < 0.05) had become newly significant in the SD. Additionally, some of the ORs from the SD had been perturbed so much that its confidence interval no longer contained the OR derived from the OD, and other ORs had shifted effects.

The number of overlapping, shifted, and newly significant effects generally increased in multivariable regression compared to univariable regression. Table 1 summarises the number of perturbations in the ORs observed from univariable and multivariable regressions, in terms of overlap or shifted OR estimates in the SD relative to the OD, and the number of statistically significant variables in the OD and SD.

**Table 1:** Summary of the number of overlapping, shifted and statistically significant associations (after one-hot encoding) comparing a synthetic dataset of 3100 subjects to the original dataset of 3735 subjects.

| Architecture |  | Overlapping<br>associations | Shifted<br>associations | Significant<br>in SD | Significant<br>in OD |
| --- | --- | --- | --- | --- | --- |
| Fast ML | Univariable | 16 / 37 | 3 / 37 | 18 / 37 | 25 / 37 |
|  | Multivariable | 26 / 37 | 1 / 37 | 14 / 37 | 18 / 37 |
| Gaussian<br>Copula | Univariable | 13 / 37 | 11 / 37 | 17 / 37 | 25 / 37 |
|  | Multivariable | 18 / 37 | 9 / 37 | 12 / 37 | 18 / 37 |
| CTGAN | Univariable | 17 / 37 | 6 / 37 | 31 / 37 | 25 / 37 |
|  | Multivariable | 20 / 37 | 7 / 37 | 13 / 37 | 18 / 37 |
| DP-CGAN | Univariable | 22 / 37 | 5 / 37 | 26 / 37 | 25 / 37 |
|  | Multivariable | 23 / 37 | 9 / 37 | 14 / 37 | 18 / 37 |

The adjusted c-statistics for multiple regression over all variables on negative body image were 0.87, 0.88, 0.86, and 0.86, for Fast ML, Gaussian Copula, CTGAN, and DP-CGAN generators, respectively.

Forest plots of the ORs (see supplementary material S6 to S13) show that the effect estimates from the OD had not been consistently reproduced in the SD, irrespective of generator architecture.

### Privacy concealment

For a given training sample, as one generates ever-larger sample sizes of SD, for all generators *except* DP-CGAN, there comes a point where a ‘real’ subject’s information becomes replicated into the SD. This would correspond to a minimum Hamming distance in given SD equalling to zero.

Figure 3 illustrates how the probability of a real patient being replicated in an SD dataset generally tends to increase for all architectures except for DP-CGAN. The figure shows the proportion of identical subjects in training and SD set, normalised to the size of the training set. The DP-CGAN appears entirely insensitive to training and generation sample size, as we were consistently able to generate up to 39100 SD samples without replicating any real subjects.

**Figure 3:**
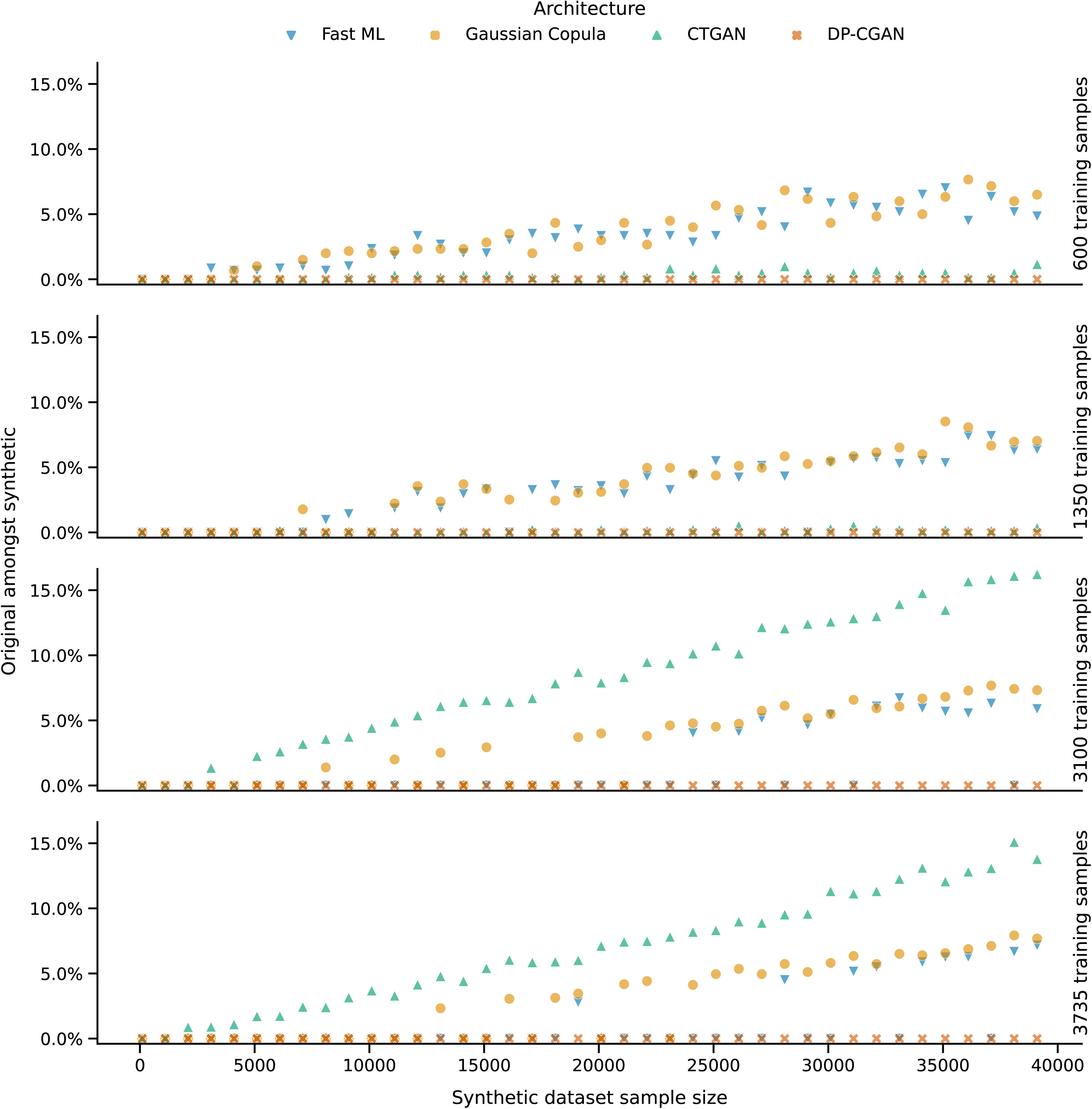
Identity concealment in relation to varying training and synthetic data sample size, per colour-coded generator architecture. Each horizontal row depicts the respective scores per training dataset sample size.

For Fast ML and Gaussian Copula, when generating 39100 subjects in the SD, there will be approximately 5% of synthetic subjects perfectly resembling ‘real’ subjects, and this occurred independently of the training sample size. The number of replicated subjects in the SD decreases roughly linearly towards zero when (obviously) the SD size is zero.

The training sample size of the CTGAN is notable; for 1100 training subjects and fewer, the number of replicated subjects is very low, barely just above that of the DP-CGAN which was always zero. However, when a training sample size of 3100 or more is used, the CTGAN switched to a state where it was much more likely than Fast ML or Gaussian Copula to replicate a real subject in the SD. This consistently reaches up to 15% of synthetic subjects perfectly resembling ‘real’ subjects, when 39100 synthetic subjects are generated. Note that this is the same range of training sample dependence where the veracity metric of density – and precision – switches from poor (∼0.50) to acceptable (∼0.70) for the CTGAN, in Figure 2. However, the training and generation loss curves (see supplementary material S14 and S15) did not specifically indicate a clear sign of ‘model collapse’ for the CTGAN in this range of training sample sizes.

Even if real subjects’ values are not fully replicated in the SD, it might still be possible to identify sensitive information through attribute inference attacks. Figure 4 plots the attribute inference score from the CAP algorithm. Higher scores imply better concealment, therefore presenting lower risk of exposing sensitive information (here, self-perception of attractiveness) via an attribute inference attack using only the SD. Concealment score for Fast ML and Gaussian Copula were insensitive to training sample size and to SD sample size; median scores were 0.41 and 0.46, respectively. Concealment in CTGAN and DP-CGAN was dependent on training sample size, but not dependent on SD output size; range of scores were 0.32 to 0.40 and 0.34 to 0.42, respectively. However, Figure 4 shows that attribute inference is not directly related to replication of real subjects in the SD.

**Figure 4:**
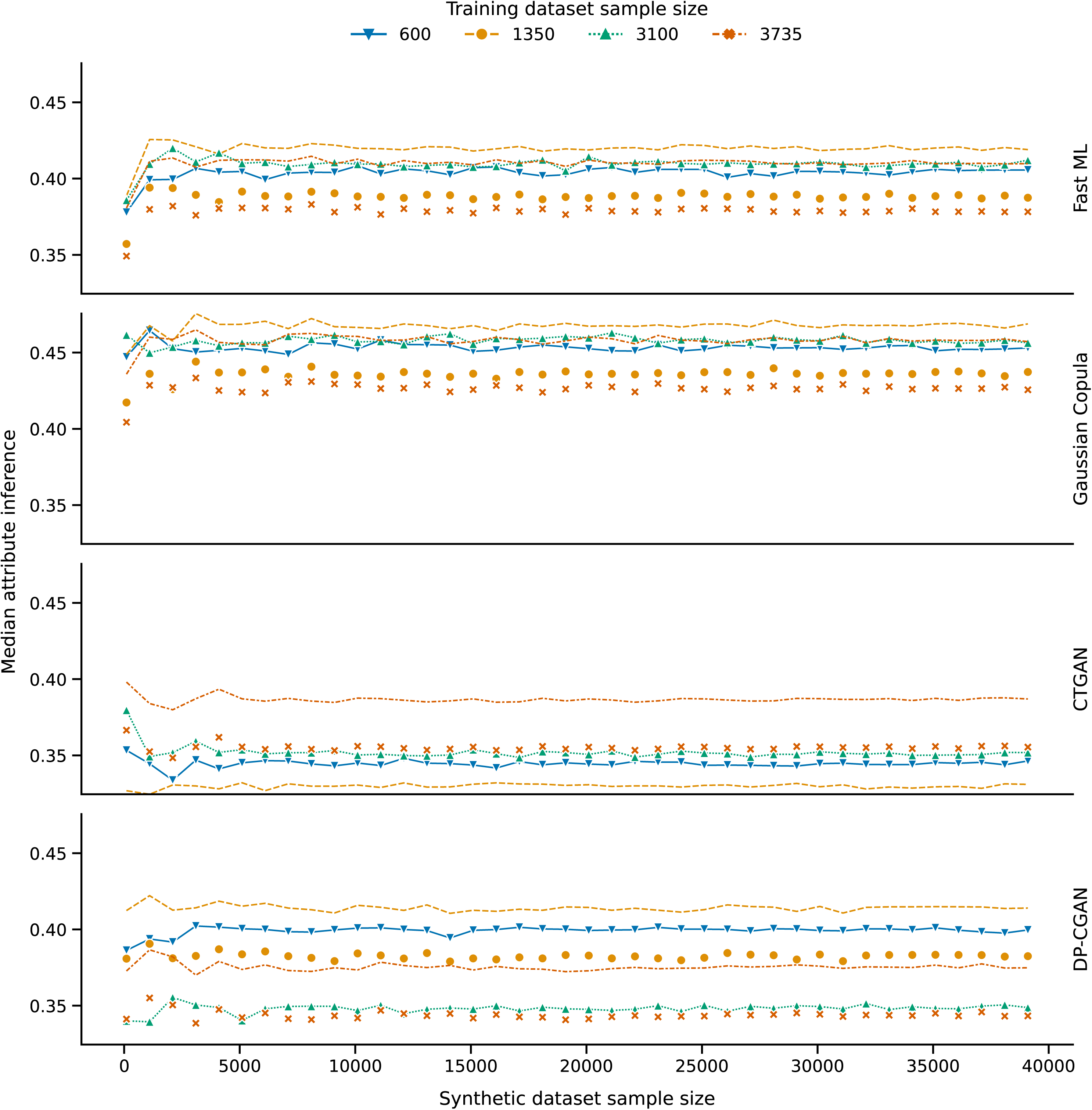
Attribute concealment in relation to varying synthetic data sample size, per colour-coded training data sample size. The median attribute inference score relates to the SDMetric’s CAP-algorithm outcome, where a value of 1 is best, and 0 is worst. Each horizontal row depicts the respective scores of the included generator architectures.

## Discussion

Sample size played an important role in SD actionability. Training and SD sample size influenced veracity, as high diversity was only obtained through an appropriate balance between training and SD sample size regardless of generator architecture; the CTGAN architecture did however require proportionally larger SD sizes to obtain equal diversity. Utility was only assessed with a SD sample size similar to the training data, and we found that inter-variable associations were not uniformly reproducible in the SD. Privacy concealment was sensitive to sample size. The Fast ML and Gaussian Copula architectures had a distinct decrease in identity concealment that was proportional to both training and synthetic sample sizes. CTGAN was able to achieve up to a factor 65 SD enlargement with smaller training datasets, but was susceptible to identity disclosure when exceeding a certain training sample sizes. The DP-CGAN architecture was the best performer in terms of identity concealment as in none of our experiments it had included any training data in the SD. All SD had inferable attributes, irrespective of generator architecture, which carried finite risk of disclosing sensitive real information.

Arora and Arora already investigated SD sample size and found it less influential, but their sample size ranges were narrow, and they acknowledged the need for broader investigation^15^. To the best of our knowledge, our combination of varying training and SD size is unique, whereas other work consistently focusses on balancing representativeness and privacy for a given training and SD sample size, hindering further comparison to our findings. The minimum Hamming distance for identity concealment indeed presents the most conservative scenario. However, the experiment was informative insofar as demonstrating the increasing risk of disclosure as SD size increases, and the susceptibility thereof among generator architectures. Inescapably, failed identity concealment can potentially result in re-identification with an unfortunate combination of identifiable attributes that act as ‘real’ data held by a bad actor^36^.

We assume most clinical SD creators have limited computational resources and know-how to perform architecture adaptation and exhaustive tuning. Thus, we chose four accessible and easy-to-use generator architectures with default settings. It is however conceivable that actionability would improve after adaptation and tuning^37^ – despite apparent over or under-fitting – yet we conceive this to be feasible only for a fraction of creators.

The implications can differ per use-case and accordingly require appropriate considerations. For instance, when aiming to augment a dataset, certain architectures appear to have an ‘upper’ enlargement limit, but also when sharing SD with a small sample size, for instance to prototype certain applications, the SD should at least be sufficiently large to capture the ‘real’ diversity. Thus, it is advisable that SD creators consistently scrutinise the suitability of their product for a certain purpose.

The (in-)dispensability of consistent SD scrutiny – in light of sample size – might substantially be influenced by ‘real’ subjects’ characteristics and sample size’s role requires further evaluation. In part, the necessity of consistent scrutiny reiterates the need for established methods to assess SD’s actionability^2,23,24^. Regardless, SD can technically be made actionable, and exuberant innovation can not only simplify this process and appraisal thereof but can incorporate here unmentioned domains such as SD fairness^38^, yet for true SD actionability in healthcare established ethical and legal consensus is paramount as these should dictate what actionability actually implies^5,8^.

Overall, SD is indeed able to resemble ‘real’ data to a moderate degree. In our experiments with AYA cancer data, we found that sample size has various roles in defining SD’s actionability. Typically, SD sample size had to be sufficiently large to encapsulate the entire veracity, yet SD sample size also should not be too large, as it might exacerbate flaws in utility and increases privacy risks. The training sample size dictated this balance between veracity, privacy, and SD sample size, in which smaller training sample sizes generally appear best suited with smaller SD sample sizes.

## Data Availability

All data produced and used in the presented work are available upon reasonable request to the authors. The code and software versioning used in the presented work are available on GitHub with a working example, see: https://github.com/MaastrichtU-CDS/AYA-synthetic-data.

## Prior presentation

This study and its results are original and have not yet been presented elsewhere.

## Acknowledgements

All individuals that have contributed to this study are included in the author list. Experiments were made possible using the Data Science Research Infrastructure (https://dsri.maastrichtuniversity.nl/) hosted at Maastricht University.

## Code availability

The code and software versioning used for this work are available on GitHub with a working example, see: https://github.com/MaastrichtU-CDS/AYA-synthetic-data.

---

**Supplementary material S1:**
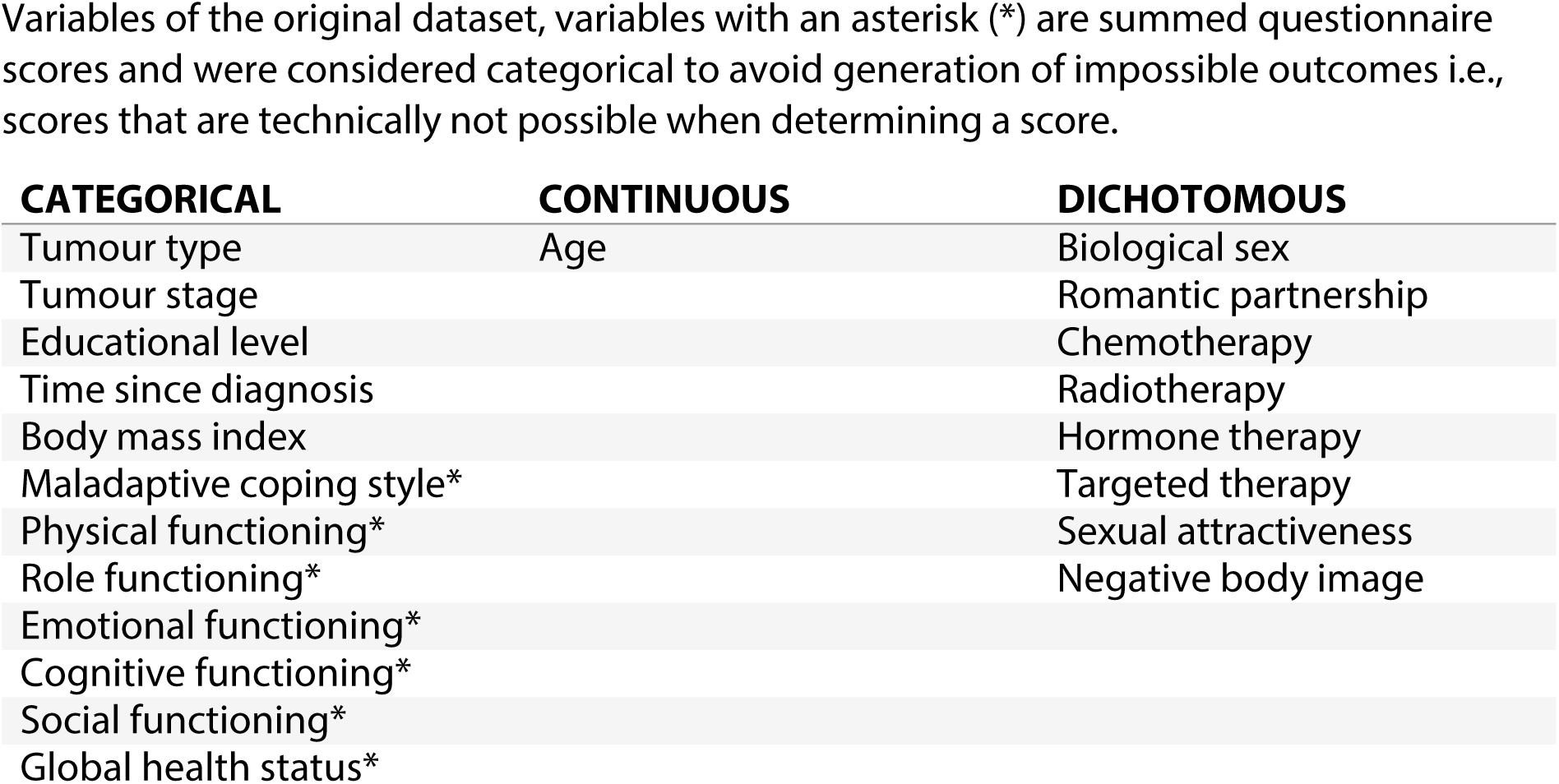
Variables of the original dataset and their datatype

**Supplementary material S2:**
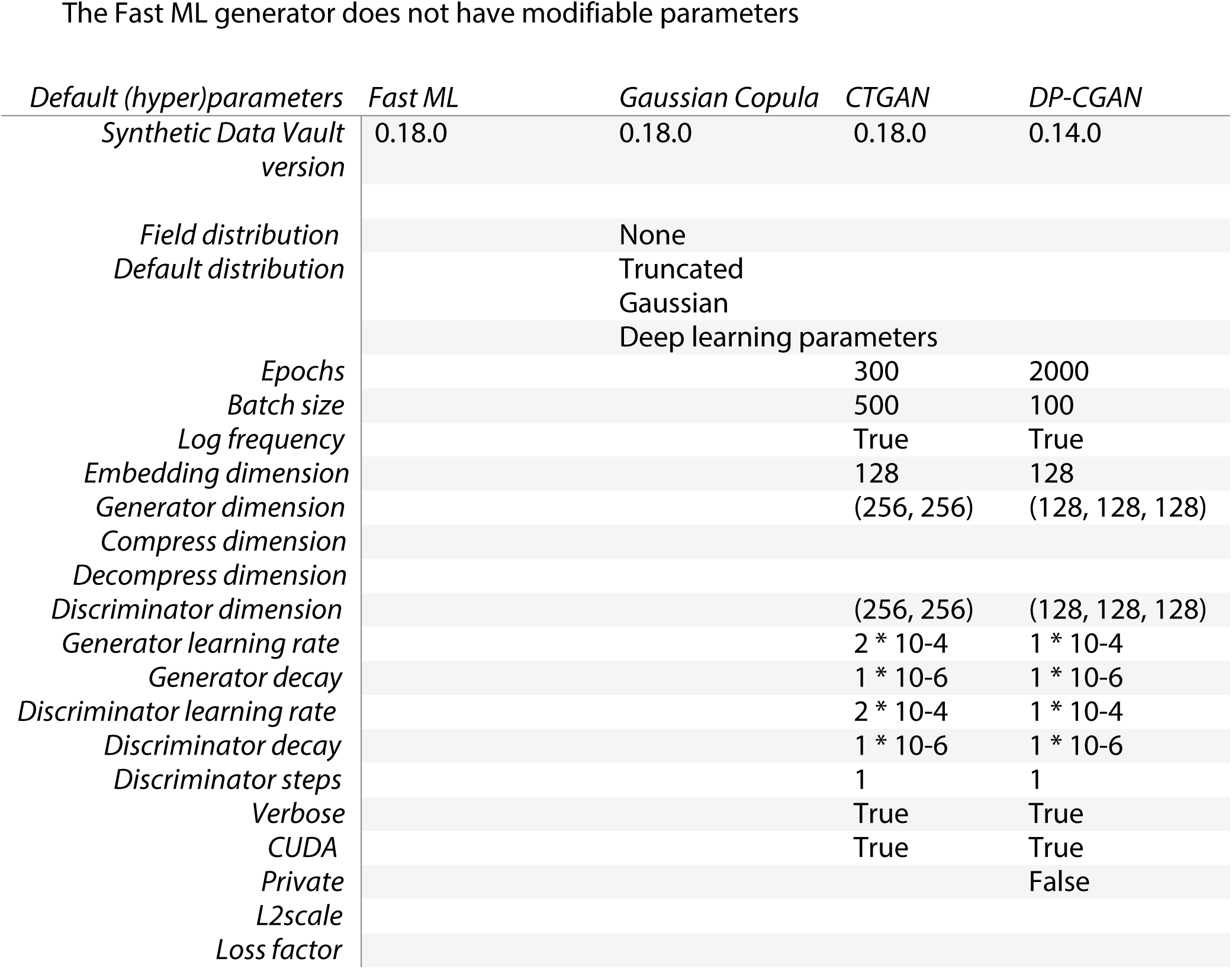
Generator (hyper-) parameters per architecture

**Supplementary material S3:**
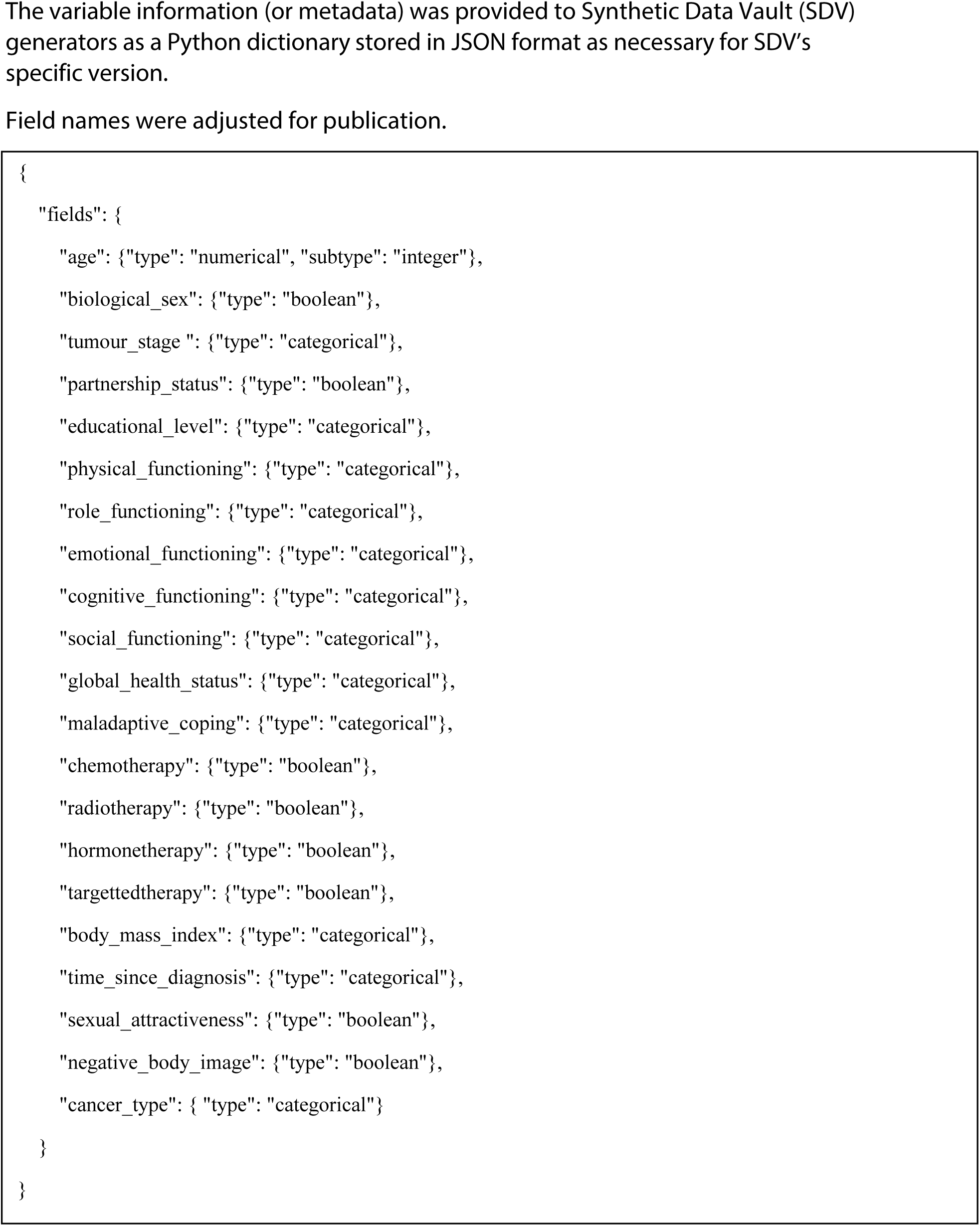
Variable metadata as provided to the generator

**Supplementary material S4:**
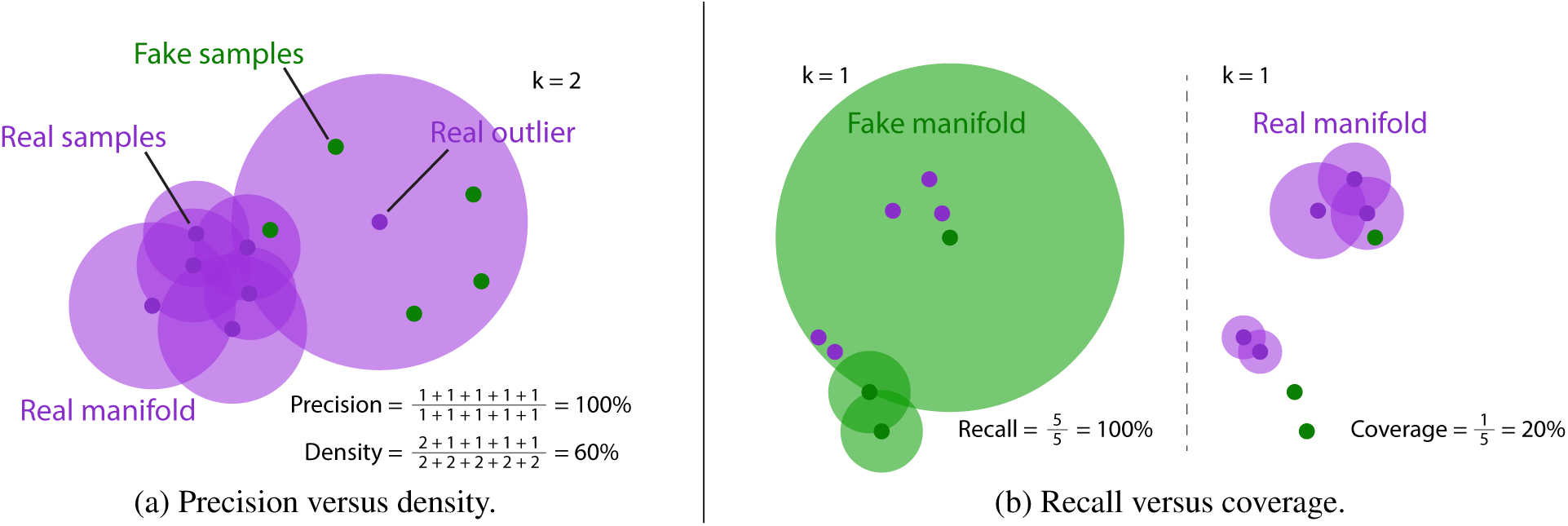
Overview of metrics. Two example scenarios for illustrating the advantage of density over precision and coverage over recall. Note that for recall versus coverage figure, the real and fake samples are identical across left and right. [Taken from the original article (DOI: 10.5555/3524938.3525603) with consent of the corresponding author].

**Supplementary material S5.**
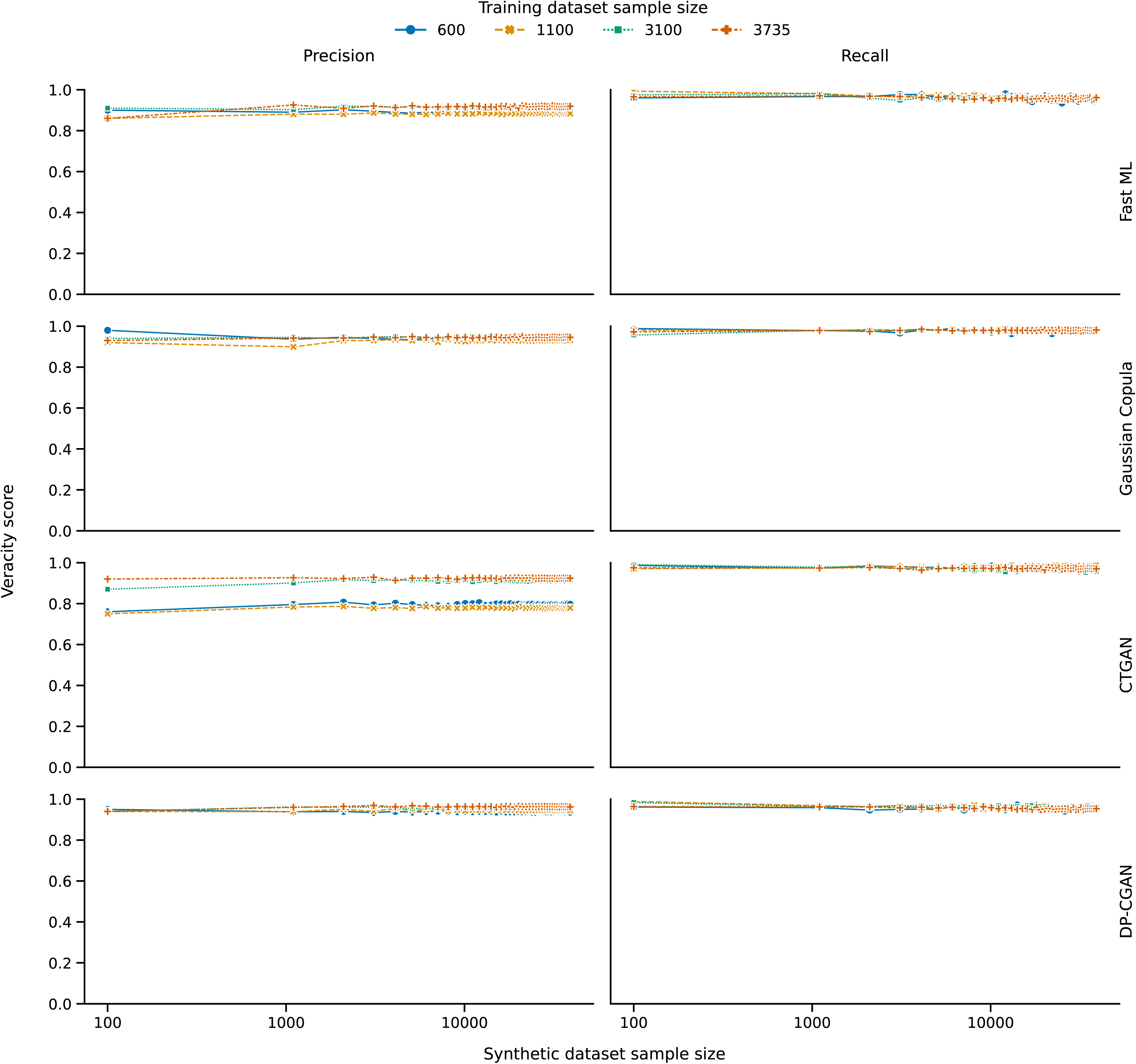

**Supplementary Material S6:**
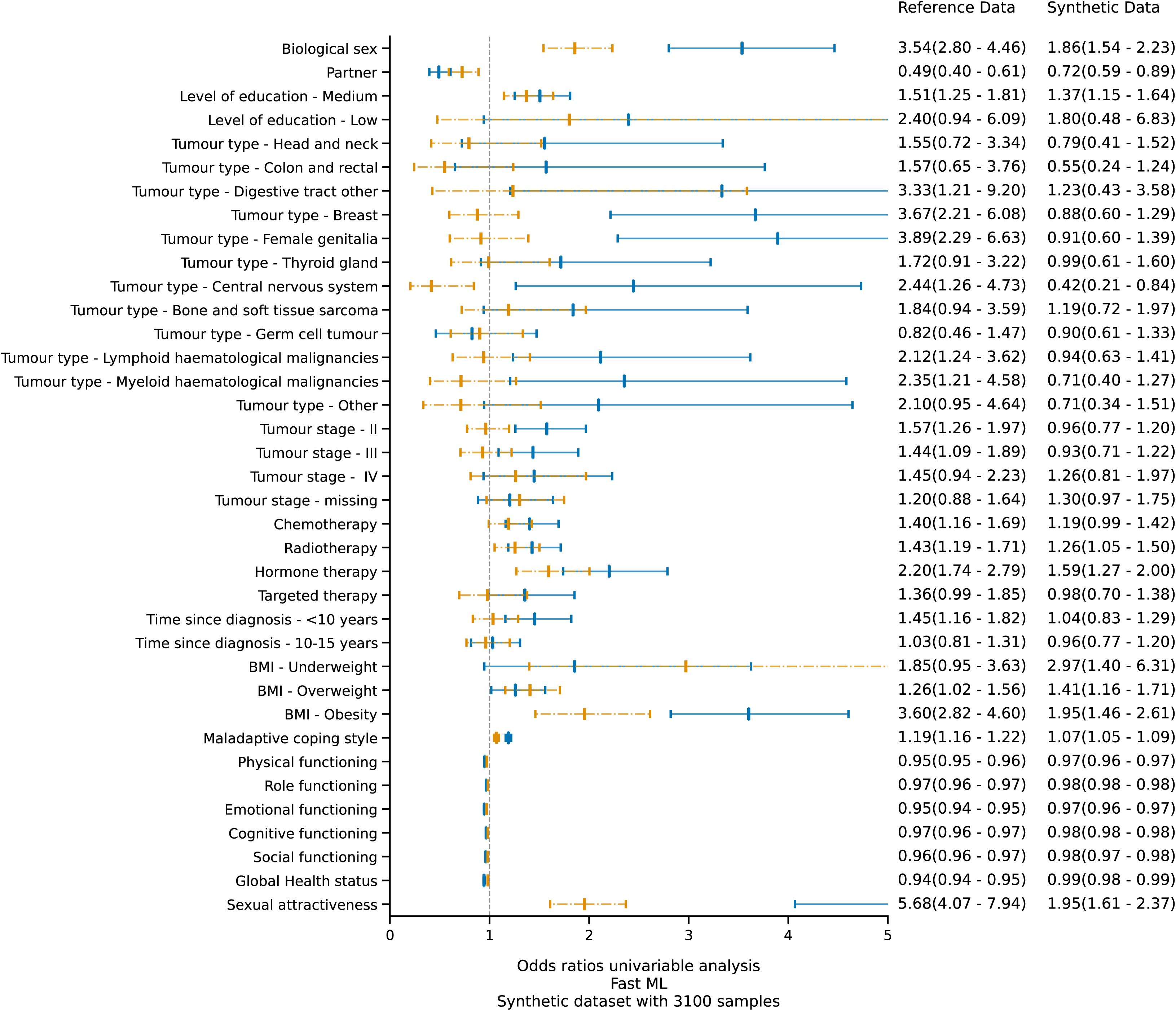
Effect measure plot for the univariable logistic regression analysis on negative body image using datasets produced by Fast ML. The blue graphics represent the odds ratios and confidence intervals of the original (training) data, whilst the orange graphics represents those belonging to the synthetic data.

**Supplementary Material S7:**
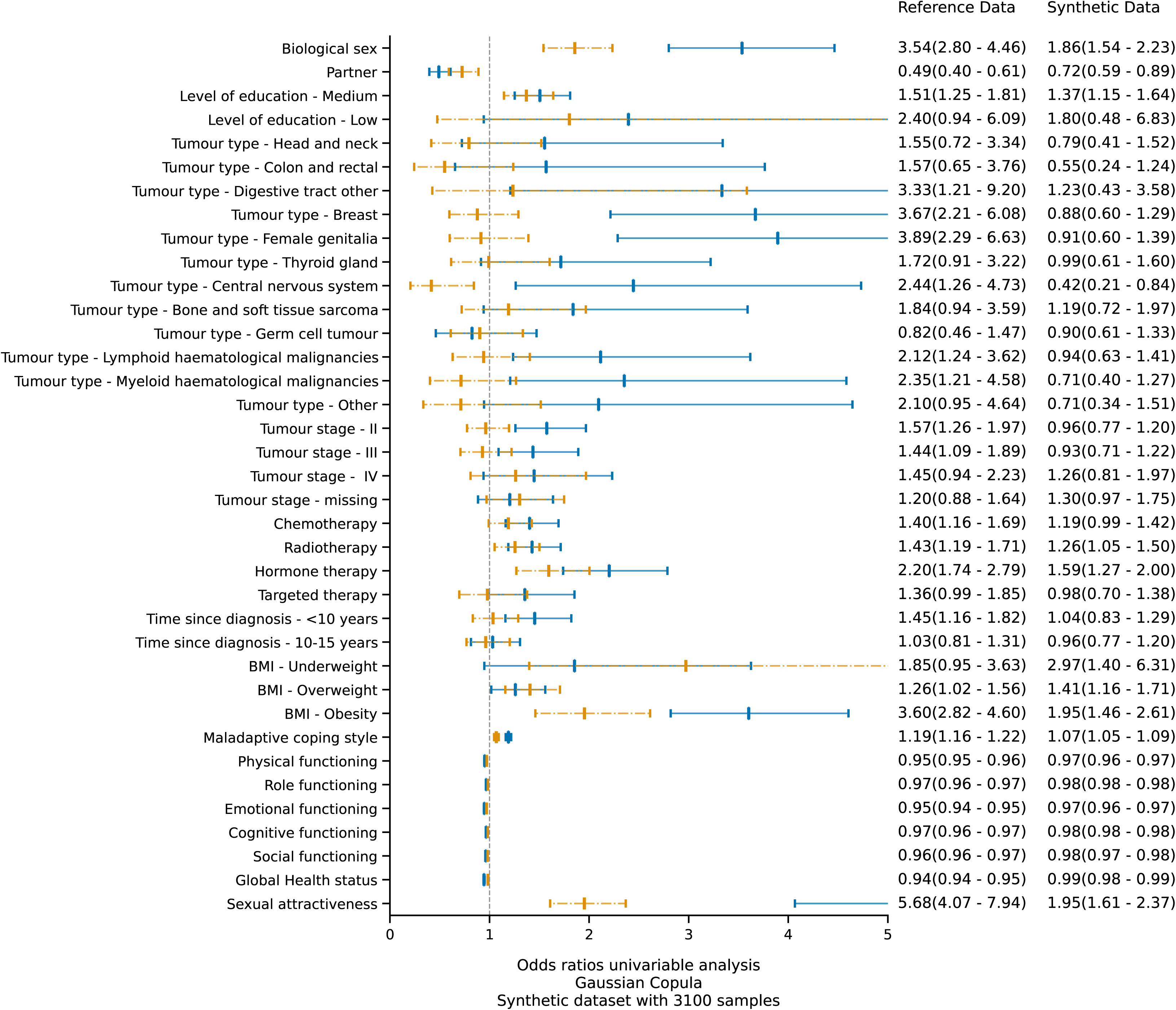
Effect measure plot for the univariable logistic regression analysis on negative body image using datasets produced by Gaussian Copula. The blue graphics represent the odds ratios and confidence intervals of the original (training) data, whilst the orange graphics represents those belonging to the synthetic data.

**Supplementary Material S8:**
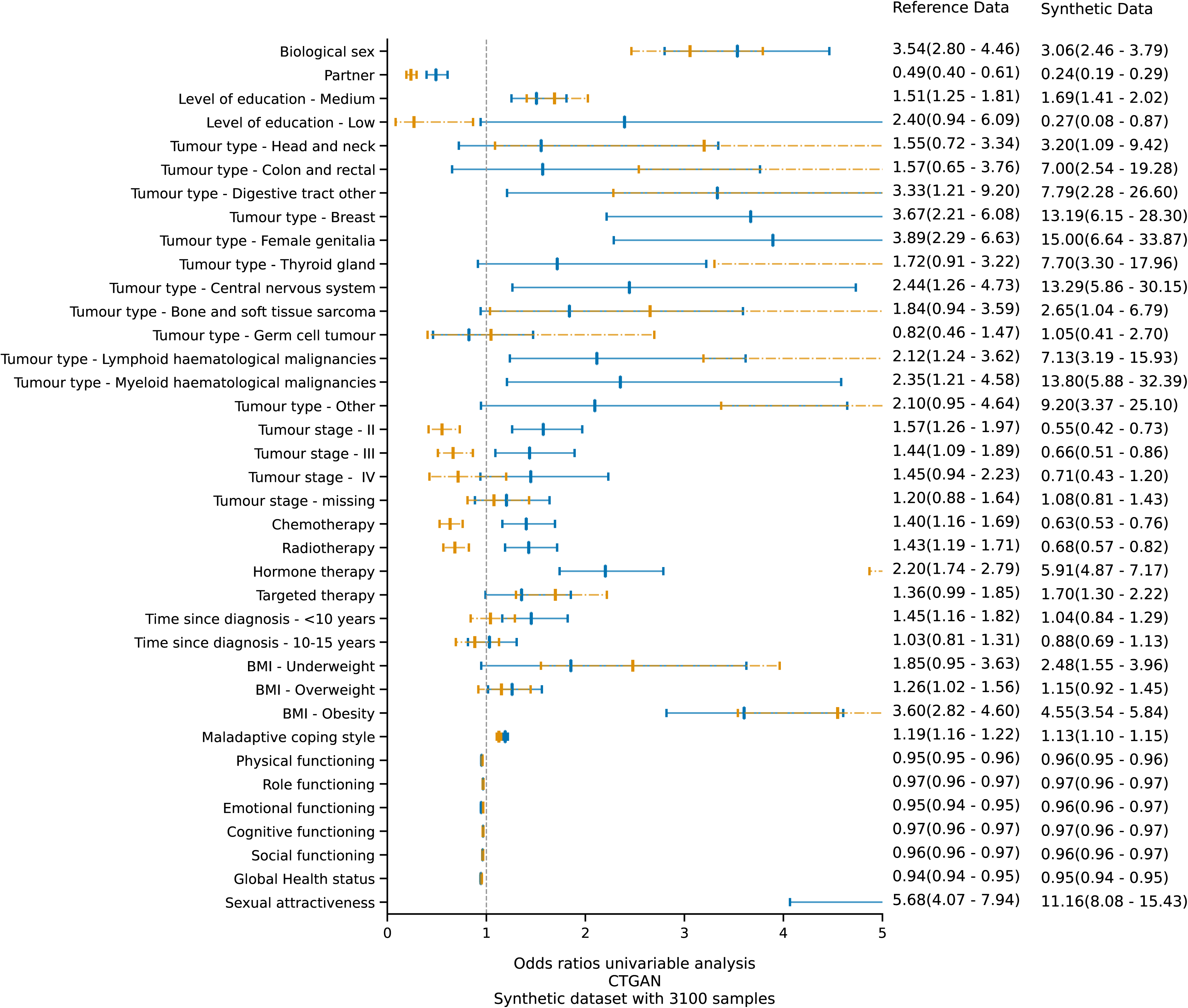
Effect measure plot for the univariable logistic regression analysis on negative body image using datasets produced by CTGAN. The blue graphics represent the odds ratios and confidence intervals of the original (training) data, whilst the orange graphics represents those belonging to the synthetic data.

**Supplementary Material S9:**
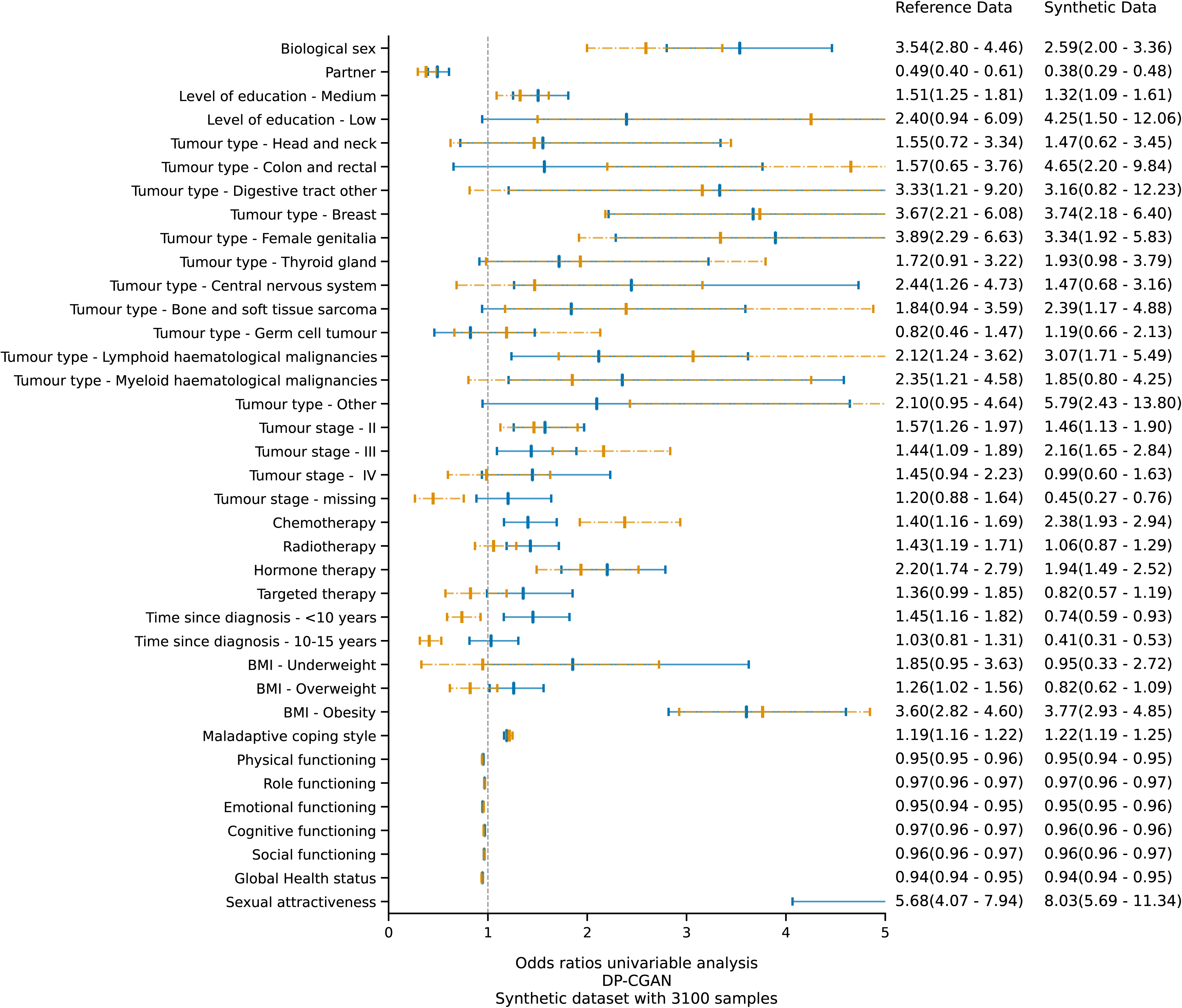
Effect measure plot for the univariable logistic regression analysis on negative body image using datasets produced by DP-CGAN. The blue graphics represent the odds ratios and confidence intervals of the original (training) data, whilst the orange graphics represents those belonging to the synthetic data.

**Supplementary Material S10:**
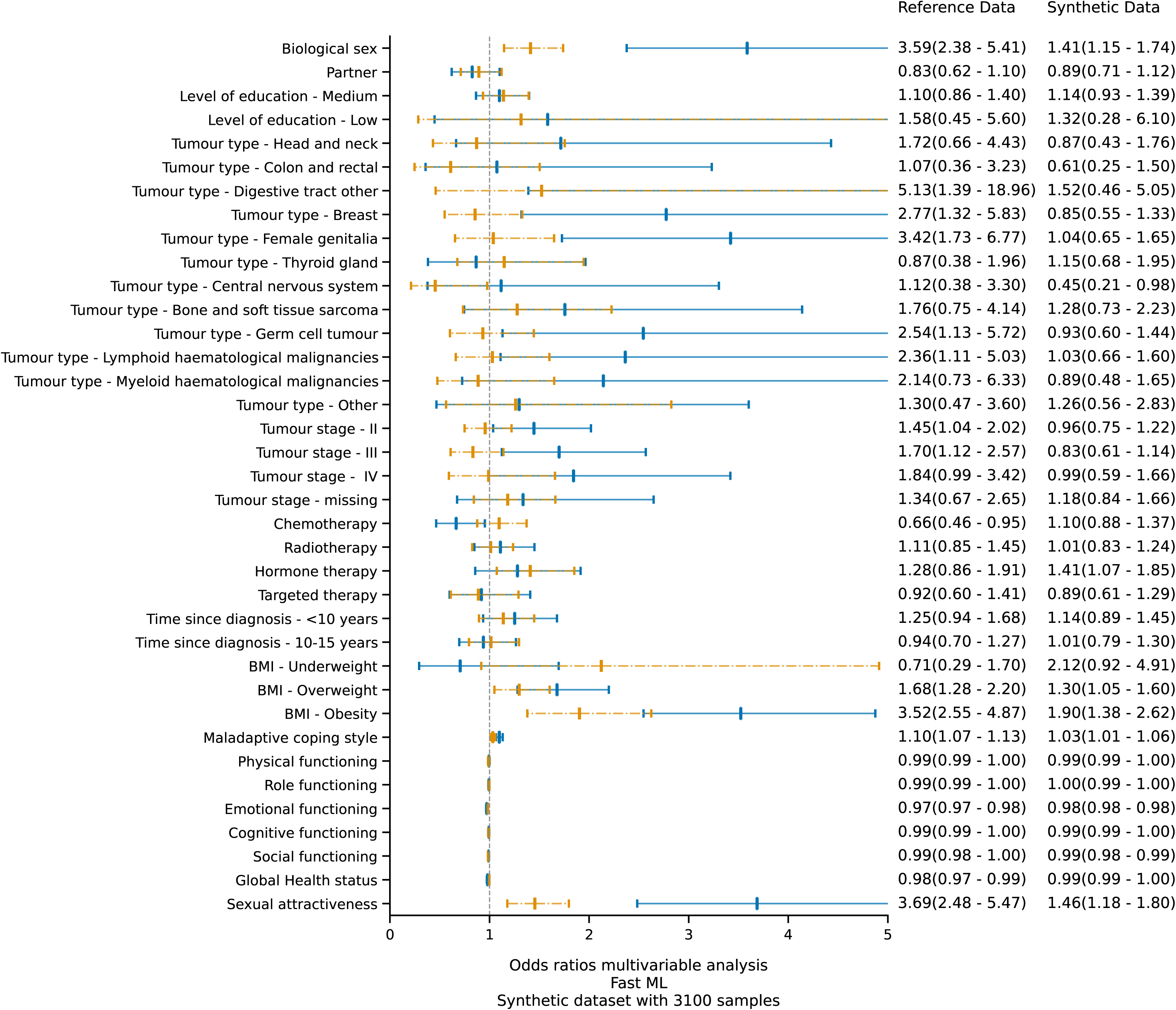
Effect measure plot for the multivariable logistic regression analysis on negative body image using datasets produced by Fast ML. The blue graphics represent the odds ratios and confidence intervals of the original (training) data, whilst the orange graphics represents those belonging to the synthetic data.

**Supplementary Material S11:**
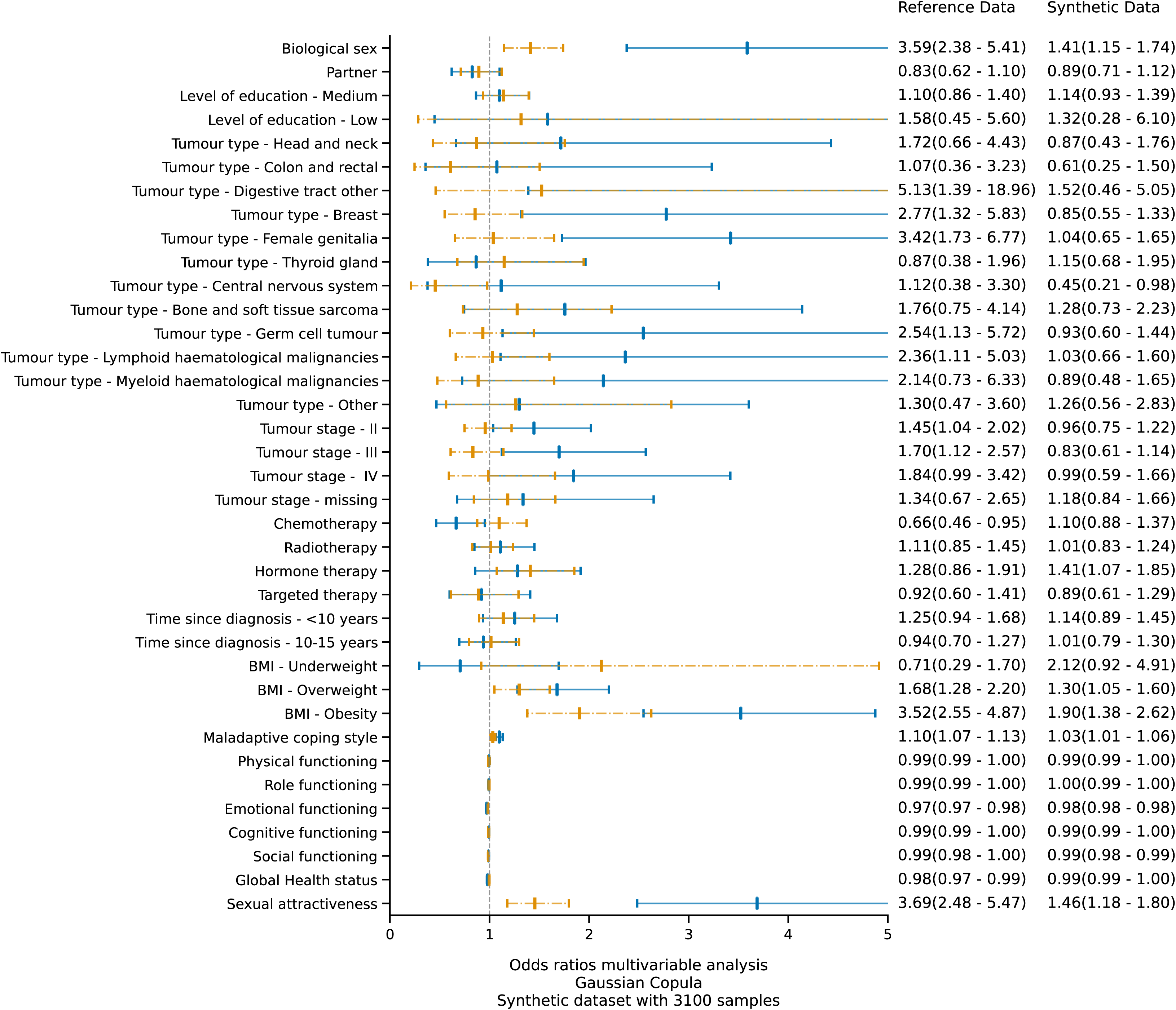
Effect measure plot for the multivariable logistic regression analysis on negative body image using datasets produced by Gaussian Copula. The blue graphics represent the odds ratios and confidence intervals of the original (training) data, whilst the orange graphics represents those belonging to the synthetic data.

**Supplementary Material S12:**
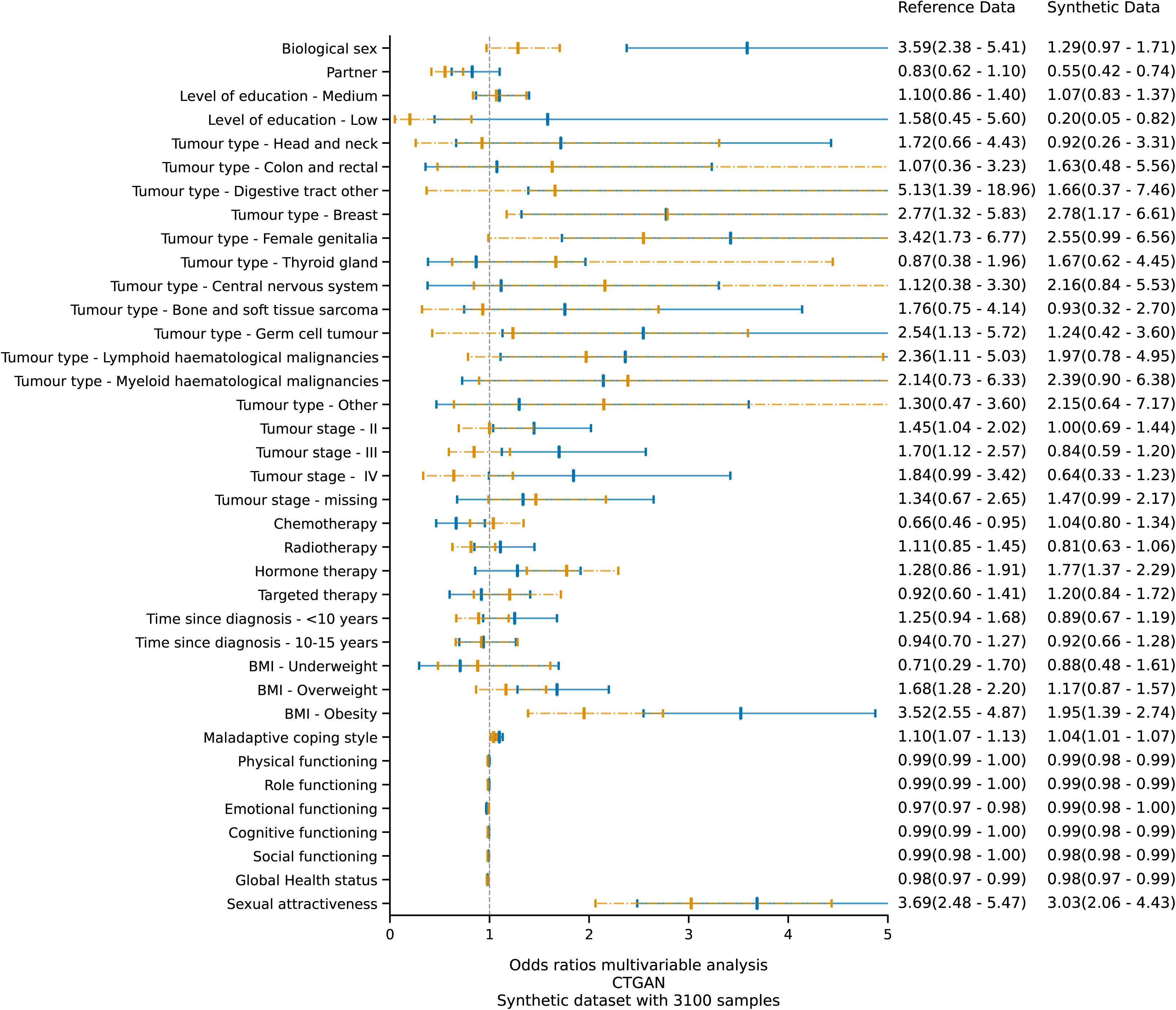
Effect measure plot for the multivariable logistic regression analysis on negative body image using datasets produced by CTGAN. The blue graphics represent the odds ratios and confidence intervals of the original (training) data, whilst the orange graphics represents those belonging to the synthetic data.

**Supplementary Material S13:**
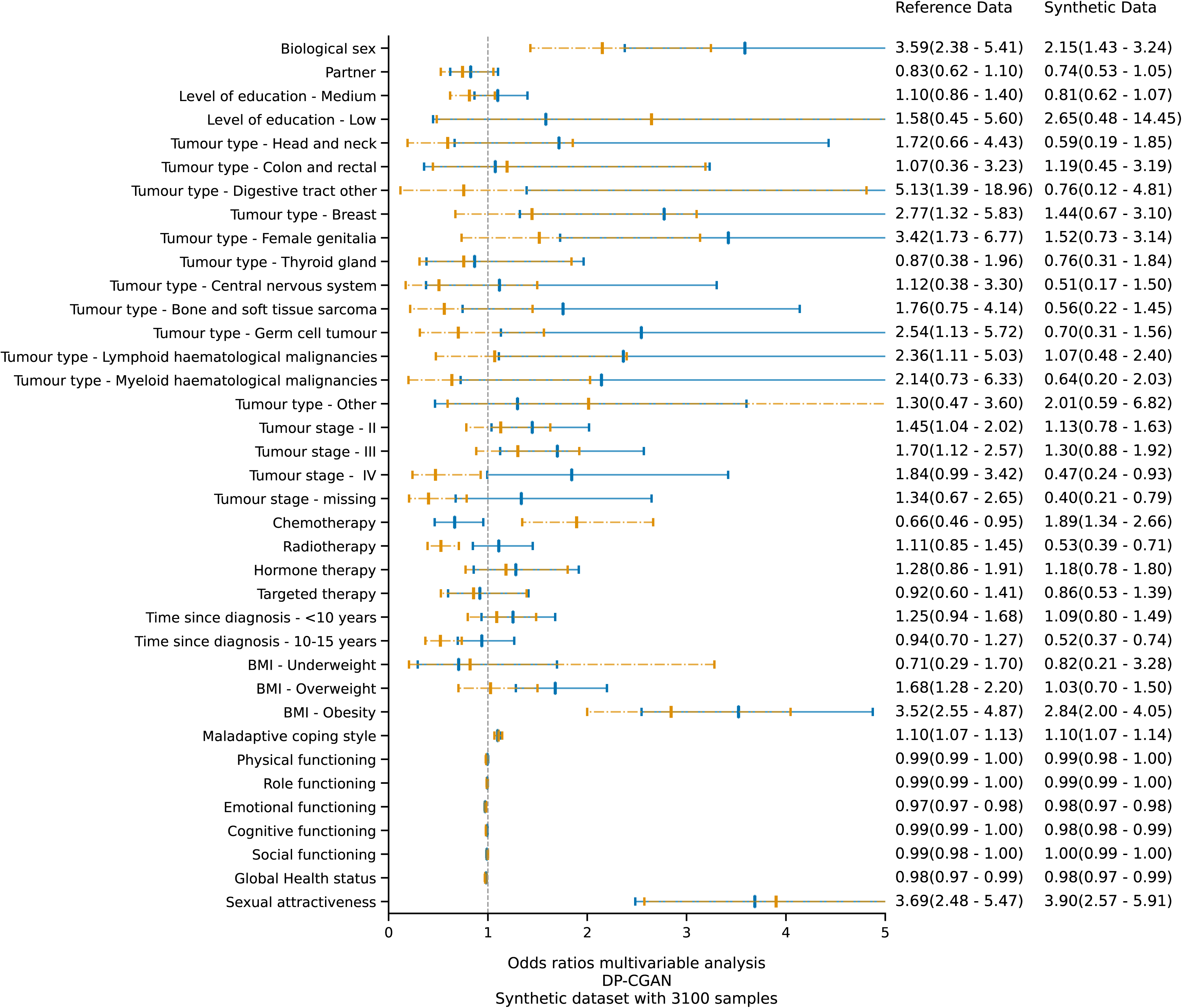
Effect measure plot for the multivariable logistic regression analysis on negative body image using datasets produced by DP-CGAN. The blue graphics represent the odds ratios and confidence intervals of the original (training) data, whilst the orange graphics represents those belonging to the synthetic data.

**Supplementary material S14:**
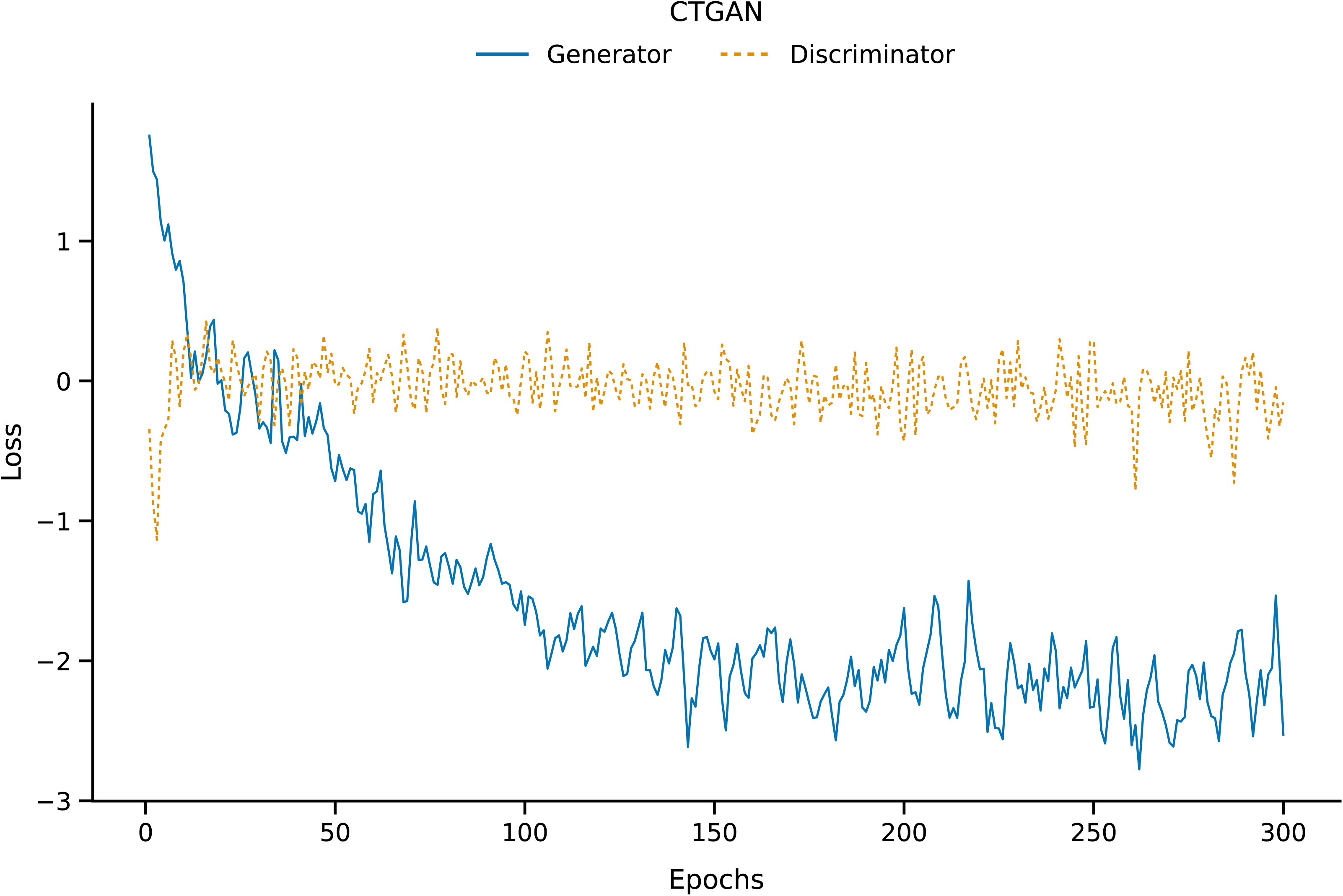
Generator and discriminator loss of a CTGAN architecture with default hyper-parameters trained on 3735 OD samples.

**Supplementary material S15:**
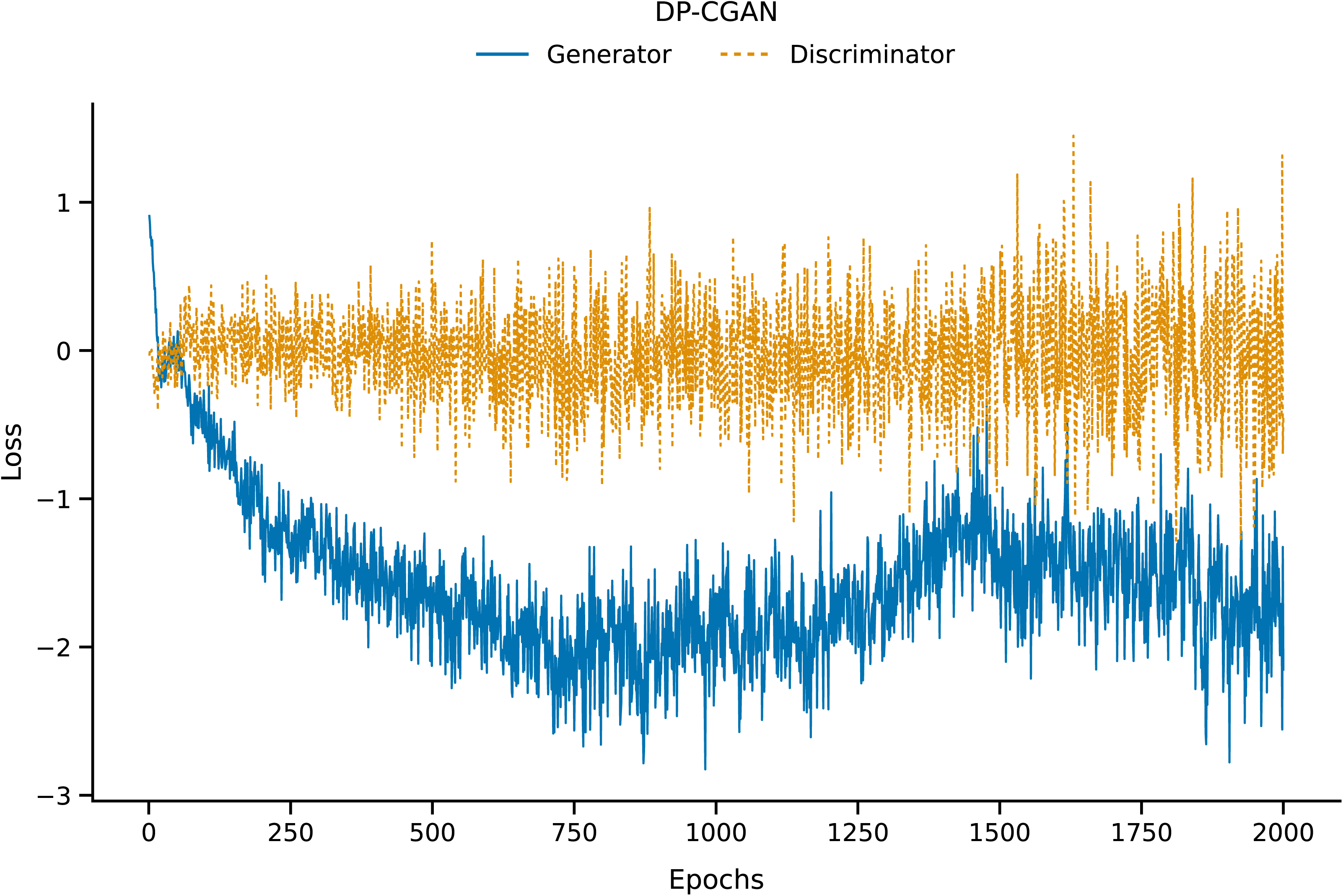
Generator and discriminator loss of a DP-CGAN architecture with default hyper-parameters trained on 3735 OD samples.

